# The reproduction number of COVID-19 and its correlation with public health interventions

**DOI:** 10.1101/2020.05.01.20088047

**Authors:** Kevin Linka, Mathias Peirlinck, Ellen Kuhl

## Abstract

Throughout the past six months, no number has dominated the public media more persistently than the reproduction number of COVID-19. This powerful but simple concept is widely used by the public media, scientists, and political decision makers to explain and justify political strategies to control the COVID-19 pandemic. Here we explore the effectiveness of political interventions using the reproduction number of COVID-19 across Europe. We propose a dynamic SEIR epidemiology model with a time-varying reproduction number, which we identify using machine learning. During the early outbreak, the basic reproduction number was 4.22±1.69, with maximum values of 6.33 and 5.88 in Germany and the Netherlands. By May 10, 2020, it dropped to 0.67±0.18, with minimum values of 0.37 and 0.28 in Hungary and Slovakia. We found a strong correlation between passenger air travel, driving, walking, and transit mobility and the effective reproduction number with a time delay of 17.24±2.00 days. Our new dynamic SEIR model provides the flexibility to simulate various outbreak control and exit strategies to inform political decision making and identify safe solutions in the benefit of global health.

## 1 Motivation

Since the beginning of the new coronavirus pandemic in December 2020, no other number has been discussed more controversially than the reproduction number of COVID-19 [36]. Epidemiologists use the basic reproduction number *R*_0_ to quantify how many new infections a single infectious individual creates in an otherwise completely susceptible population [13]. The public media, scientists, and political decision makers across the globe have started to adopted the basic reproduction number as an illustrative metric to explain and justify the need for community mitigation strategies and political interventions [21]: An outbreak will continue for *R*_0_ > 1 and come to an end for *R*_0_ < 1 [25]. While the concept of *R*_0_ seems fairly simple, the reported basic reproduction number for COVID-19 varies hugely depending on country, culture, calculation, stage of the outbreak [36]. Knowing the precise number of *R*_0_ is important, but challenging, because of limited data and incomplete reporting [12]. It is difficult–if not impossible–to measure *R*_0_ directly [50]. The earliest COVID-19 study that followed the first 425 cases of the Wuhan outbreak via direct contact tracing reported a basic reproduction number of 2.2 [33]. However, especially during the early stages of the outbreak, information was limited because of insufficient testing, changes in case definitions, and overwhelmed healthcare systems [47]. Most basic reproduction numbers of COVID-19 we see in the public media today are estimates of mathematical models that depend critically on the choice of the model, the initial conditions, and numerous other modeling assumptions [12]. To no surprise, the mathematically predicted basic reproduction numbers cover a wide range, from 2.2–3.6 for exponential growth models to 4.1–6.5 for more sophisticated compartment models [36].

Compartment models are a popular approach to simulate the epidemiology of an infectious disease [29]. A prominent compartment model is the SEIR model that represents the timeline of a disease through the interplay of four compartments that contain the susceptible, exposed, infectious, and recovered populations [6]. The SEIR model has three characteristic parameters, the transition rates *β* from the susceptible to the exposed state, *α* from the exposed to the infectious state, and *γ* from the infectious to the recovered state [25]. The latter two are disease specific parameters associated with the inverses of the latent period *A* = 1/*α* during which an individual is exposed but not yet infectious, and the infectious period *C* = 1/*γ* during which an individual can infect others [32]. For COVID-19, depending on the way of reporting, these two times can vary anywhere between *A* = 2 to 6 days and *C* = 3 to 18 days [40, 42, 44]. The most critical feature of any epidemiology model is the transition from the susceptible to the exposed state. This transition typically scales with the size of the susceptible and infectious populations *S* and *I*, and with the contact rate *β*, the inverse of the contact period *B* = 1/*β* between two individuals of these populations [25]. The product of the infectious period and the contact rate defines the reproduction number *R* = *Cβ* [12]. Community mitigation strategies and political interventions seek to reduce the contact rate *β*, and with it the reproduction number *R*, to control the outbreak of a pandemic [44].

The first official case of COVID-19 in Europe was reported on January 24, 2020. Within only 45 days, the pandemic spread across all 27 countries of the European Union [15]. On March 17, for the first time in its history, the European Union closed all its external borders to prevent a further spreading of the disease [16]. Within the following two weeks, many local governments supplemented the European regulations with lockdowns and national travel restrictions. In response, passenger air travel within the European Union dropped by up to 95% [18]. These drastic measures have stimulated a wave of criticism, especially because initially, it was entirely unclear to which extent they would succeed in reducing the number of new infections [38].

In this study, as Europe begins to relax these constraints, we correlate the effect of Europe-wide travel restrictions to the outbreak dynamics of COVID-19. We introduce a dynamic SEIR model with a time-varying contact rate *β* (*t*) that transitions smoothly from the initial contact rate *β*_0_ at the beginning of the outbreak to the effective contact rate *β*_t_ under global travel restrictions and local lockdown. We express the time-varying contact rate *β* (*t*) = *R*(*t*)/*C* as a function of the effective reproduction number *R*(*t*) and use Bayesian inference to learn the evolution of the reproduction number for each country of the European Union from its individual outbreak history [15]. Our model allows us to precisely quantify the initial basic reproduction number *R*_0_, the effective reproduction number *R*_t_, and the adaptation time *t*^∗^ to achieve this reduction, which are important quantitative metrics of the effectiveness of national public health intervention. Our model also specifies the exact time delay *Δt* between the implementation of political actions and their effects on the outbreak dynamics of COVID-19. This time delay is particularly important to plan exit strategies and estimate risks associated with gradually or radically relaxing current local lockdowns and global travel restrictions.

## 2 Methods

### Epidemiology modeling

We model the epidemiology of the COVID-19 outbreak using an SEIR model with four compartments, the susceptible, exposed, infectious, and recovered populations, governed by a set of ordinary differential equations [34], see Appendix,

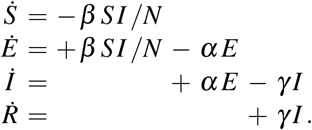

The transition rates between the four compartments, *β, α*, and *γ*, are inverses of the contact period *B* = 1/*β*, the latent period *A* = 1/*α*, and the infectious period *C* = 1/*γ*, and *N* = *S* + *E* + *I* + *R* is the total population. We interpret the latency rate *α* and the infectious rate *γ* as disease-specific for COVID-19, and assume that they are constant across all 27 countries of the European Union. We interpret the contact rate *β* = *β* (*t*) as behavior specific, and assume that it is different for each country and can vary in time to reflect the effect of societal and political actions. For easier interpretation, we express the contact rate *β* (*t*) = *R*(*t*)/*C* in terms of the time-varying effective reproduction number *R*(*t*). For the effective reproduction number, we postulate a hyperbolic tangent type ansatz,

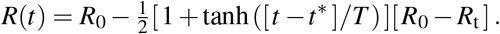

This ansatz ensures a smooth transition from the basic reproduction number *R*_0_ at the beginning of the outbreak to the current reproduction number *R*_t_ under travel restrictions and lockdown, where *t*^∗^ is the adaptation time and *T* is the transition time, see Appendix.

### COVID-19 outbreak and mobility data

We draw the COVID-19 outbreak data for all 27 countries of the European Union [15]. From these data, we extract the newly confirmed cases as the difference between today’s and yesterday’s reported cases. We sample all European air traffic data from the Eurocontrol dashboard, a pan-European Organization dedicated to support European aviation [19]. In addition, we approximate car, walking, and transit mobility using a database generated from cell phone data [4]. These data represent the relative volume of location requests per city, subregion, region, and country, scaled by the baseline volume on January 13, 2020. We smoothen the weekday-weekend fluctuations in outbreak and mobility data by applying a moving averaging window of seven days.

### Machine learning

To analyze the evolution of the effective reproduction number for each country, and predict possible exit scenarios, we identify the initial exposed and infectious populations *E*_0_ and *I*_0_ and the effective reproduction number *R*(*t*) using the reported COVID-19 cases in all 27 countries of the European Union [15]. For each country, our simulation window begins on the day at which the number of reported cases surpasses 100 individuals and ends on May 10, 2020 for the initial simulation and on June 20, 2020 for the prediction. We fix the latency and infectious periods to *A* = 2.5 days and *C* = 6.5 days [31, 33, 47]. To account for uncertainties in the initial exposed and infectious populations *E*_0_ and *I*_0_ and in the effective reproduction number *R*(*t*), we use Bayesian inference with Markov-Chain Monte-Carlo to estimate the following set of model parameters *ϑ*= {*E*_0_, *I*_0_, *σ, R*_0_, *R*_t_, *t*^∗^, *T*}. Here, *σ* represents the width of the likelihood 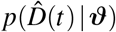 between the time-varying reported new cases 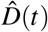 and the simulated affected population *D*(*t, ϑ*). We adopt a Student’s t-distribution for the likelihood between the data and the model predictions [11, 30] with a confirmed case number-dependent width,

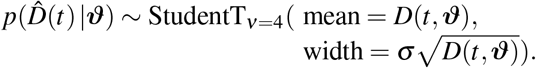

We apply Bayes’ rule to obtain the posterior distribution of the parameters [41, 45] using the prior distributions in Table 1 and the reported case numbers [15],

**Table 1.** Prior distributions for the initial exposed and infectious populations *E*_0_ and *I*_0_, width of likelihood *σ*, basic and effective reproduction numbers *R*_0_ and *R*_t_, adaptation time *t*^∗^, and transition time *T*.

| Parameter | Distribution |
| --- | --- |
| $E_0$ | LogNormal($\log(D(t=A)), 1.5$) |
| $I_0$ | LogNormal($\log(D(t=0)), 1.5$) |
| $\sigma$ | HalfCauchy($\beta = 1$) |
| $R_0$ | Normal(2.5, 2) |
| $R_t$ | Normal(2.5, 2) |
| $t^*$ | Normal(10, 10) |
| $T$ | LogNormal($\log(3), 1.5$) |

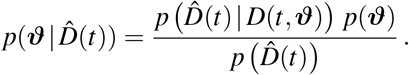

We solve this distribution numerically using the NO-U-Turn sampler [26] implementation of the python package PyMC3 [46]. We use two chains: The first 1000 samples are used to tune the sampler, and are later discarded; the subsequent 1000 samples are used to estimate the set of parameters *ϑ*. Chain convergence requires a geometric ergodicity between the Markov transition and the target distribution. In PyMC3 this is detected by split 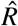 statistics, which identifies convergence by comparing the variance between the chains. From the converged posterior distributions, we sample multiple combinations of parameters that describe the time evolution of reported cases. These posterior samples allow us to quantify the uncertainty on each parameter.

To probe the effect of different exit strategies, we explore three possible projections of the effective reproduction number *R*(*t*) for each posterior parameter sample set and predict the outbreak dynamics for a 40-day period after our initial manuscript submission, from May 10 until June 20, 2020. The first scenario assumes a constant effective reproduction number *R*(*t*) = *R*_t_, the second and third scenarios simulate the effect of a linear return from *R*_t_ to the country-specific basic reproduction number *R*_0_, either rapidly within one month, or more gradually within three months. In the revision of our manuscript, we added the reported daily new cases from May 10 until June 20, 2020 to compare our model predictions against the real case data.

## 3 Results

Figure 1 illustrates the outbreak dynamics of COVID-19 for all 27 countries of the European Union. The dots represent daily new cases. The brown and red curves illustrate the fit of the SEIR model and the effective reproduction number for the time period until May 10, 2020. The gray shaded area highlights the model predictions for the 40-day period of gradual reopening, from May 10 until June 20, 2020. The dashed brown, orange, and red curves illustrate the projections for three possible exit strategies: a constant continuation at the effective reproduction number *R*_t_ from May 10, 2020, a gradual return to the basic reproduction number *R*_0_ within three months, and a rapid to *R*_0_ within one months.

**Fig 1.**
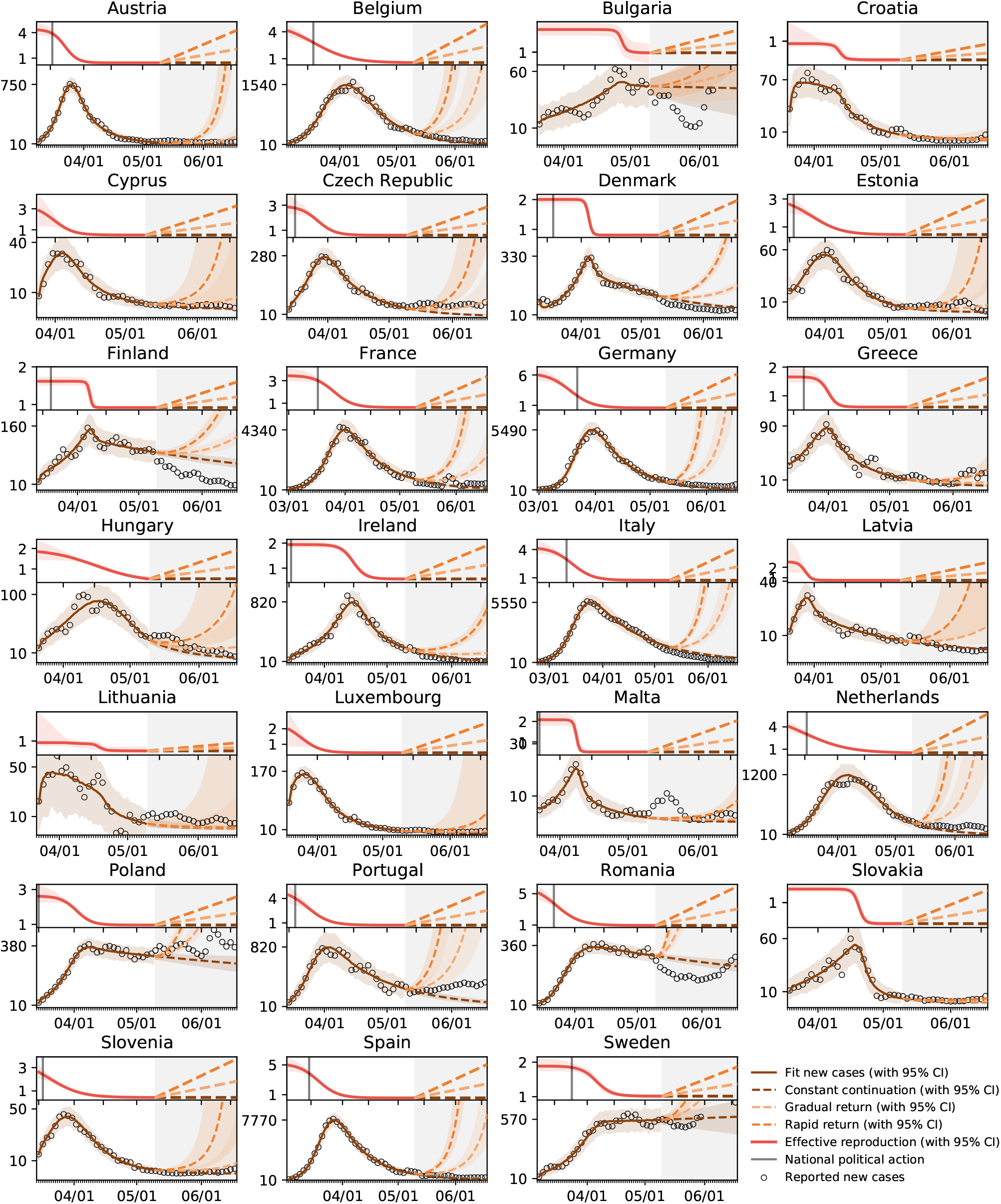
Outbreak dynamics of COVID-19 across Europe and prediction of different exit strategies. The dots represent daily new cases. The brown and red curves illustrate the fit of the SEIR model and the effective reproduction number for the time period until May 10, 2020. The gray shaded area highlights the model predictions from May 10 until June 20, 2020. The dashed brown, orange, and red curves illustrate the projections for three possible exit strategies: a constant continuation at the effective reproduction number *R*_t_ from May 10, 2020, a gradual return to the basic reproduction number *R*_0_ within three months, and a rapid to *R*_0_ within one months.

Table 2 and Figures 2 and 3 summarize the basic reproduction number *R*_0_ at the beginning of the COVID-19 out-break and the effective reproduction number *R*_t_ as of May 10, 2020. The basic reproduction number *R*_0_ has maximum values in Germany, the Netherlands, and Spain, with 6.33, 5.88, and 5.19 and minimum values in Bulgaria, Croatia, and Lithuania with 1.29, 0.93, and 0.91. The population weighted mean of the basic reproduction number across the European Union is *R*_0_ = 4.22±1.69. The effective reproduction number *R*_t_ is significantly lower than the initial basic reproduction number *R*_0_. In most countries, it is well below the critical value of *R*_t_ = 1.0. It has maximum values in Sweden, Bulgaria, and Poland all with 1.01, 0.99, and 0.96 and minimum values in Lithuania, Hungary, and Slovakia with 0.41, 0.37, and 0.28. The population weighted mean of the basic reproduction number across the European Union is *R*_t_ = 0.67±0.18.

**Table 2.** Parameters of the COVID-19 outbreak across Europe. Basic reproduction number *R*_0_, effective reproduction number *R*_t_, adaptation time *t*^∗^, adaptation speed *T*, and time delay *Δt* for fixed latency period *A* = 2.5 days and infectious period *C* = 6.5 days.

| Country | Population | $R_0$ | $R_t$ | $t^*$ | $T$ | $\Delta t$ |
| --- | --- | --- | --- | --- | --- | --- |
| Austria | 8.840.521 | 4.38±0.36 | 0.45±0.01 | 13.37±0.68 | 6.49±0.47 | 8.33±1.70 |
| Belgium | 11.433.256 | 5.00±0.73 | 0.54±0.03 | 13.31±2.84 | 19.30±1.57 | 4.00±2.35 |
| Bulgaria | 7.025.037 | 1.29±0.04 | 0.99±0.07 | 37.04±1.99 | 1.64±1.56 | 43.00±2.83 |
| Croatia | 4.087.843 | 0.93±0.22 | 0.49±0.03 | 22.36±2.90 | 2.46±3.61 | 27.33±2.36 |
| Cyprus | 1.189.265 | 3.35±1.14 | 0.50±0.02 | 6.60±2.87 | 8.02±1.47 | 14.00±0.00 |
| Czech Republic | 10.629.928 | 2.92±0.47 | 0.60±0.01 | 14.04±1.71 | 8.44±1.12 | 14.00±1.73 |
| Denmark | 5.793.636 | 2.00±0.05 | 0.81±0.01 | 24.74±0.29 | 1.72±0.45 | 24.00±2.92 |
| Estonia | 1.321.977 | 3.12±0.78 | 0.45±0.04 | 10.72±3.80 | 14.19±2.87 | 12.50±2.06 |
| Finland | 5.515.525 | 1.62±0.05 | 0.92±0.01 | 25.05±0.51 | 1.20±0.68 | 24.25±2.49 |
| France | 66.977.107 | 3.46±0.29 | 0.62±0.02 | 24.79±1.30 | 10.58±1.17 | 10.50±1.50 |
| Germany | 82.905.782 | 6.33±0.64 | 0.58±0.01 | 17.06±1.39 | 12.41±0.71 | 3.25±1.92 |
| Greece | 10.731.726 | 1.66±0.12 | 0.61±0.02 | 18.93±0.87 | 4.38±1.22 | 17.33±3.09 |
| Hungary | 9.775.564 | 1.97±0.55 | 0.37±0.15 | 25.62±6.55 | 20.23±7.33 | 31.67±1.89 |
| Ireland | 4.867.309 | 1.94±0.06 | 0.57±0.03 | 30.78±0.53 | 5.94±1.28 | 30.00±3.46 |
| Italy | 60.421.760 | 4.25±0.42 | 0.74±0.01 | 19.24±1.57 | 12.06±1.13 | 5.00±0.71 |
| Latvia | 1.927.174 | 2.50±0.89 | 0.76±0.01 | 6.99±1.32 | 2.70±0.90 | 14.67±1.89 |
| Lithuania | 2.801.543 | 0.91±0.88 | 0.41±0.09 | 26.23±9.88 | 2.25±6.25 | 34.67±1.89 |
| Luxembourg | 607.950 | 2.42±1.21 | 0.46±0.01 | 5.77±4.20 | 8.78±1.88 | 10.00±2.35 |
| Malta | 484.630 | 2.08±0.14 | 0.51±0.03 | 16.24±0.42 | 1.21±0.51 | 23.00±0.00 |
| Netherlands | 17.231.624 | 5.88±0.88 | 0.49±0.03 | 7.61±3.12 | 23.25±1.92 | 0.75±2.77 |
| Poland | 37.974.750 | 2.62±0.26 | 0.96±0.01 | 18.15±1.47 | 7.38±1.47 | 20.33±0.94 |
| Portugal | 10.283.822 | 5.10±0.86 | 0.73±0.02 | 8.93±1.86 | 10.40±1.22 | 8.67±2.62 |
| Romania | 19.466.145 | 6.06±0.84 | 0.95±0.01 | 8.12±1.60 | 11.55±0.70 | 8.33±2.36 |
| Slovakia | 5.446.771 | 1.46±0.04 | 0.28±0.03 | 31.80±0.47 | 2.65±0.70 | 40.25±0.43 |
| Slovenia | 2.073.894 | 3.83±0.96 | 0.44±0.03 | 4.65±3.50 | 15.33±1.96 | 6.33±2.36 |
| Spain | 46.796.540 | 5.19±0.50 | 0.57±0.01 | 15.90±1.17 | 10.70±0.69 | 5.50±2.60 |
| Sweden | 10.175.214 | 1.89±0.09 | 1.01±0.03 | 29.70±1.30 | 7.99±2.65 | 23.75±2.77 |
| European Union | 446.786.293 | 4.22±1.69 | 0.67±0.18 | 18.61±6.43 | 10.82±4.65 | 17.24±2.00 |

**Fig 2.**
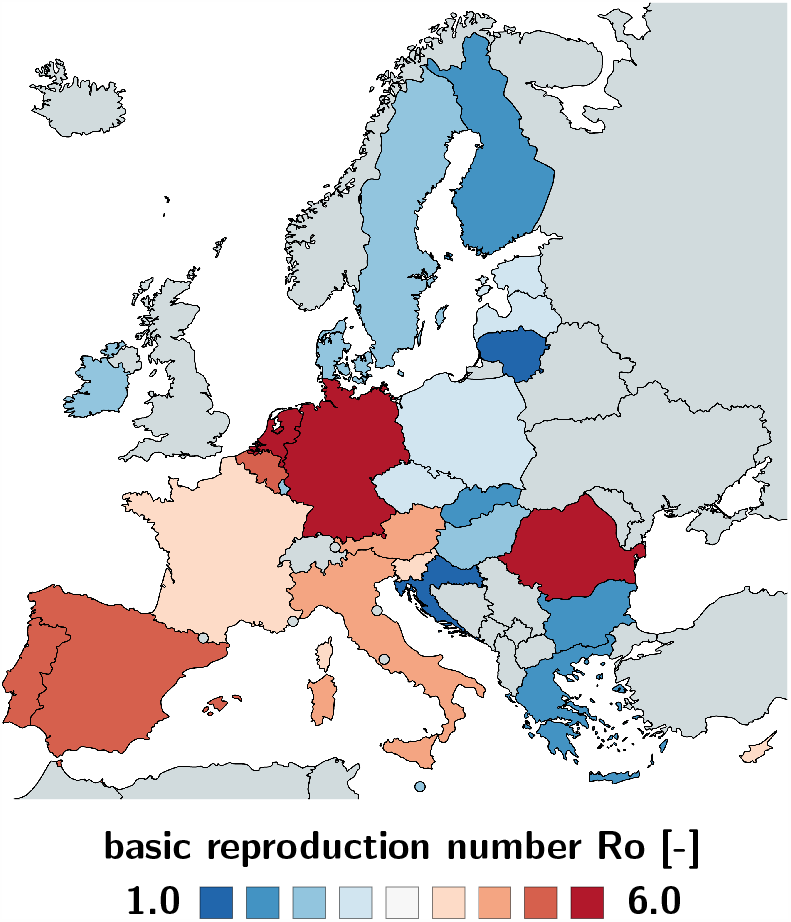
Basic reproduction number *R*_0_ of the COVID-19 outbreak across Europe. The basic reproduction number characterizes the initial number of new infectious created by one infectious individual. It has maximum values in Germany, the Netherlands, and Spain, with 6.33, 5.88, and 5.19 and minimum values in Bulgaria, Croatia, and Lithuania with 1.29, 0.93, and 0.91.

**Fig 3.**
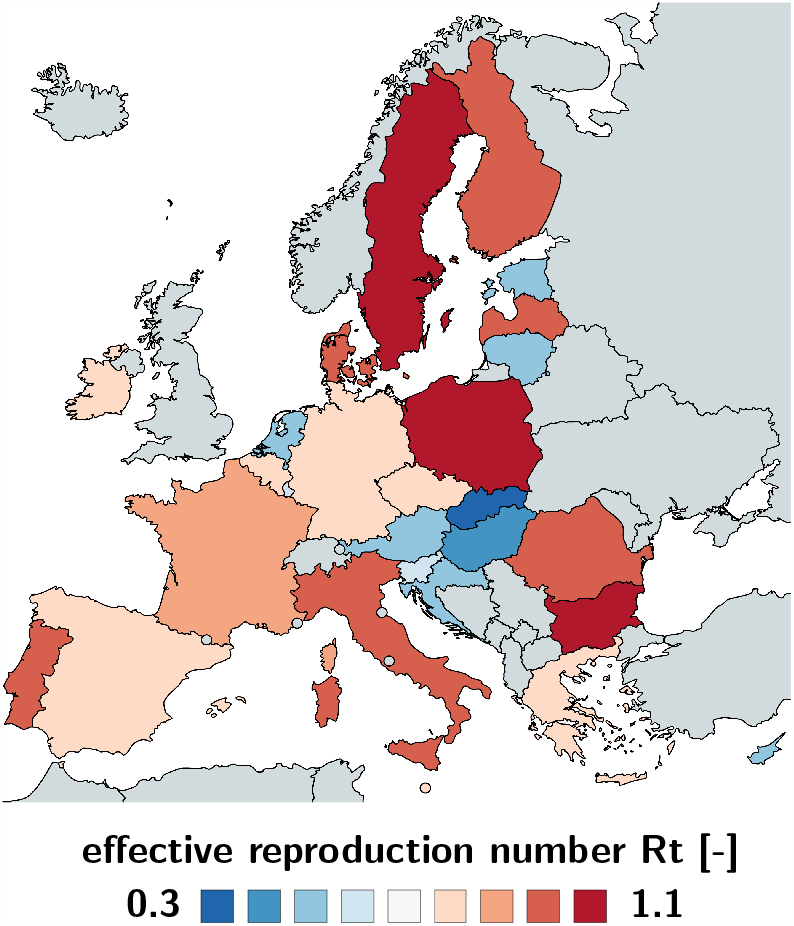
Effective reproduction number *R*_*t*_ of the COVID-19 outbreak across Europe. The effective reproduction number characterizes the current number of new infectious created by one infectious individual. It has maximum values in Sweden, Bulgaria, and Poland all with 1.01, 0.99, and 0.96 and minimum values in Lithuania, Hungary, and Slovakia with 0.41, 0.37, and 0.28 as of May 10, 2020.

Figure 4 provides a direct correlation between the reduction in mobility and the effective reproduction number of the COVID-19 outbreak across Europe. The purple, blue, grey, and black dots represent the reduction in air traffic, driving, walking, and transit mobility, the red curves show effective reproduction number with 95% confidence interval. The mean time delay *Δt* highlights the temporal delay between reduction in mobility and effective reproduction number. Spearman’s rank correlation *ρ*, a measure of the statistical dependency between both variables, reveals the strongest correlation in the Netherlands, Germany, Ireland, Spain, and Sweden with 0.99 and 0.98. Only in Slovakia, Slovenia and Lithuania, where the number of cases has not yet plateaued and the effective reproduction number does not show a clear smoothly decaying trend, there is no significant correlation between mobility and the effective reproduction number.

**Fig 4.**
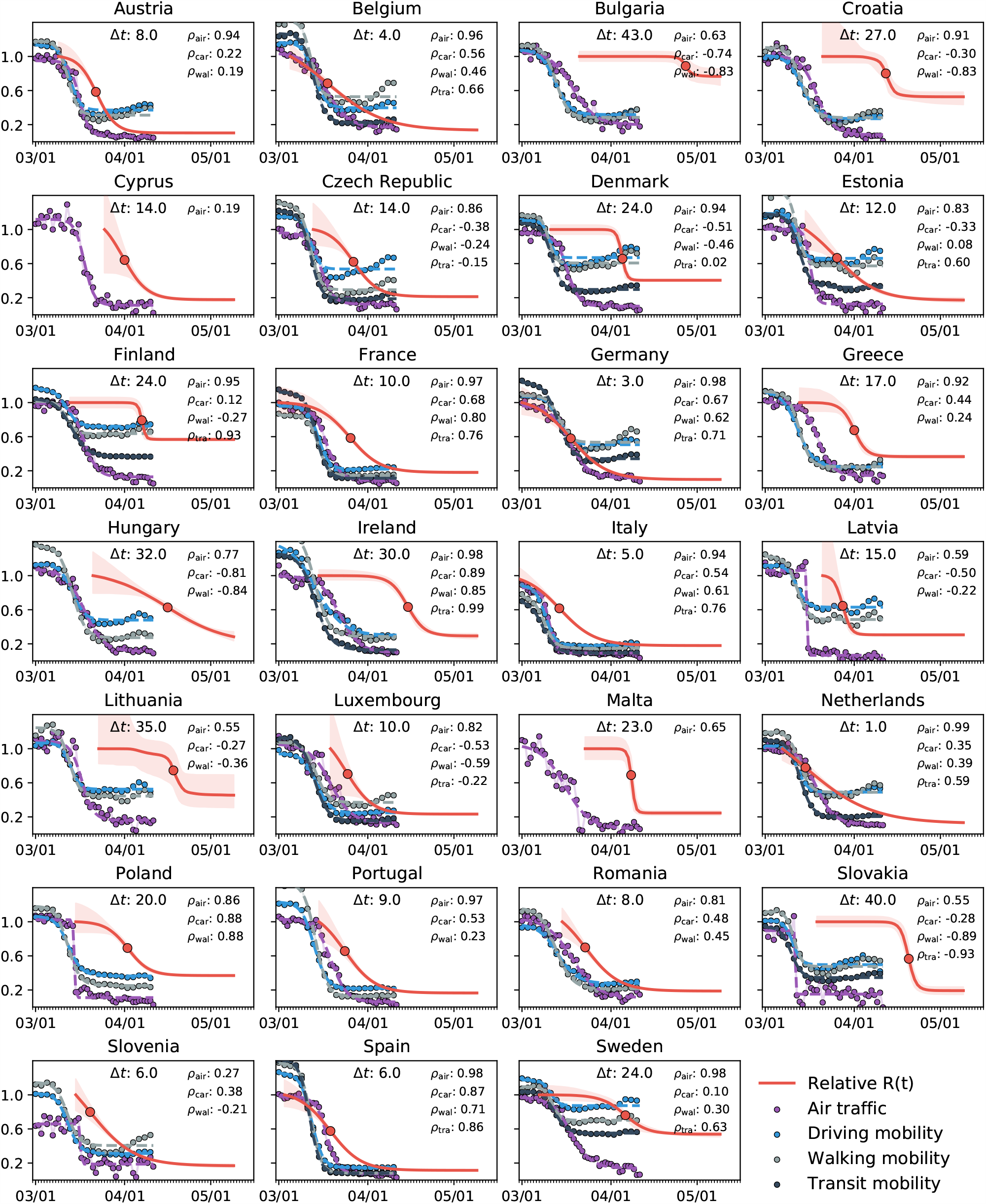
Correlation between reduction in mobility and effective reproduction number of the COVID-19 outbreak across Europe. Purple, blue, grey, and black dots represent reduction in air traffic, driving, walking, and transit mobility; red curves show effective reproduction number *R*(*t*) with 95% confidence interval. The mean time delay *Δt* highlights the temporal delay between reduction in mobility and effective reproduction number. Spearman’s rank correlation *ρ*, measures of the statistical dependency between mobility and reproduction, and reveals the strongest correlation in the Netherlands, Germany, Ireland, Spain, and Sweden with 0.99 and 0.98.

Figure 5 summarizes the learned basic reproduction number *R*_0_, the effective reproduction number *R*_t_, the adaptation time *t*^∗^, and the time delay *Δt* for all 27 countries of the European Union. The adaptation time *t*∗ characterizes the time between the beginning of the outbreak at 100 confirmed cases and the reduction in the effective reproduction number and is a quantitative measure for the reaction time in the population. The time delay Δ*t* characterizes the mean time between the reduction in air travel, driving, walking, and transit mobility and the reduction in the effective reproduction number and is a quantitative measure for the effect of mobility.

**Fig 5.**
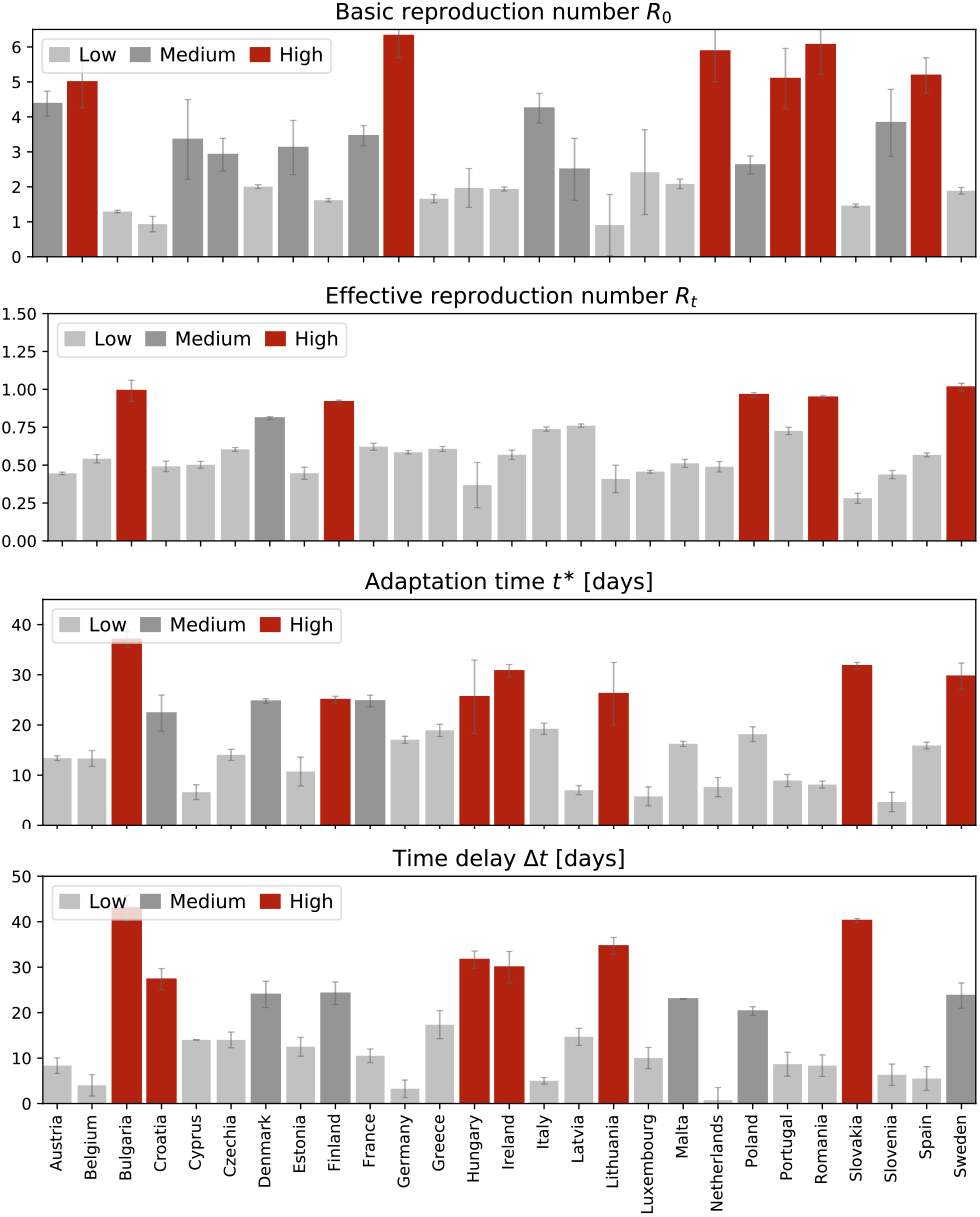
Parameters of the COVID-19 outbreak across Europe. Basic reproduction number *R*_0_, effective reproduction number *R*_*t*_, adaptation time *t*\* and time delay *Δt*. The adaptation time *t*\* characterizes the time between the beginning of the outbreak and the reduction in the effective reproduction number; the time delay *Δt* characterizes the mean time between the reduction in air travel, driving, walking, and transit mobility and the reduction in the effective reproduction number.

Table 2 and Figures 6 and 7 summarize the adaptation time *t*^∗^ and the time delay *Δt*. The adaptation time *t*^∗^ has maximum values in Bulgaria and Slovakia with 37.04 and 31.80 days and minimum values in Luxembourg and Slovenia with 5.77 and 5.64 days. The mean adaptation time across the European Union is *t*^∗^ = 18.61±6.43 days. The time delay *Δt* has maximum values in Bulgaria and Slovakia with 43.00 and 40.25 days and minimum values in Germany and the Netherlands both with 3.25 and 0.75 days. The mean time delay across the European Union is *Δt* = 17.24±2.00 days.

**Fig 6.**
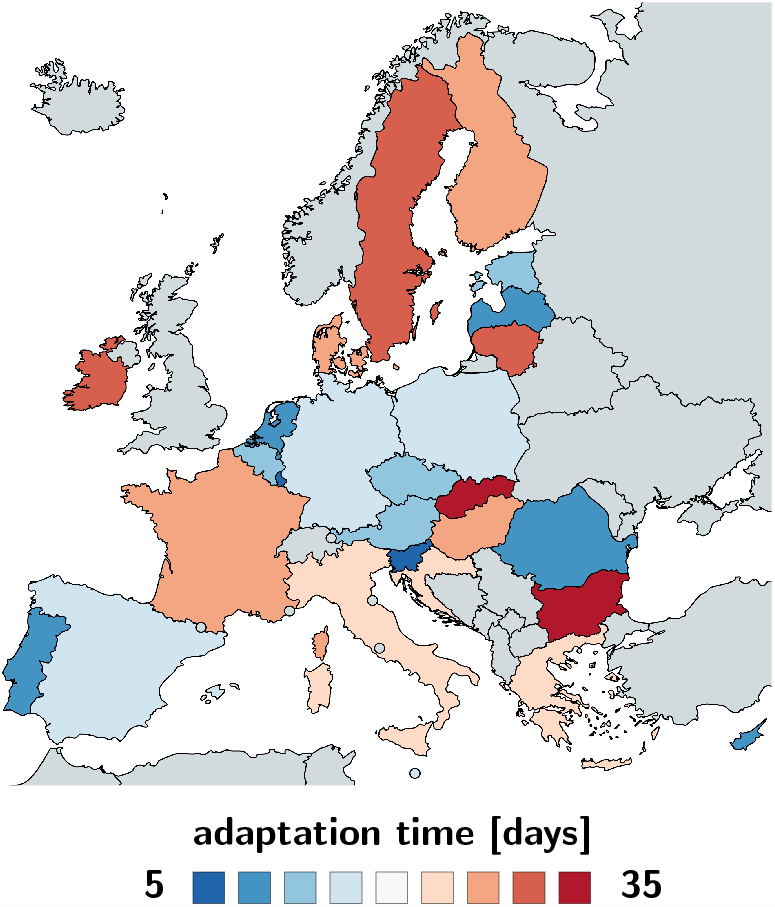
Adaptation time *t*\* between beginning of the outbreak and reduction of the effective reproduction number across Europe. The adaptation time characterizes the time between the beginning of the outbreak at 100 confirmed cases and the reduction in the effective reproduction number. It has maximum values in Bulgaria and Slovakia with 37.04 and 31.80 days and minimum values in Luxembourg and Slovenia with 5.77 and 5.64 days.

**Fig 7.**
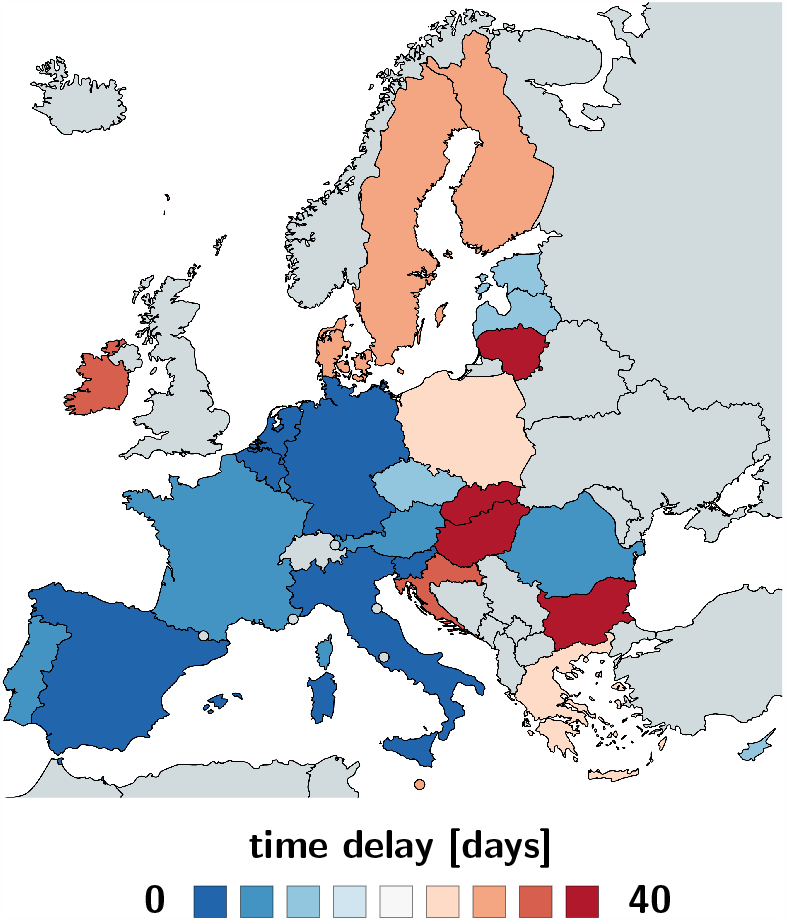
Time delay *Δt* between reduction of air travel and reduction of the effective reproduction number across Europe. The time delay characterizes the mean time between the reduction in air travel, driving, walking, and transit mobility and the reduction in the effective reproduction number. It has maximum values in Bulgaria and Slovakia with 43.00 and 40.25 days and minimum values in Germany and the Netherlands both with 3.25 and 0.75 days.

## 4 Discussion

### Mathematical models can inform political interventions

As many countries begin to explore safe exit strategies from total lockdown, shelter in place, and national travel restrictions to manage the COVID-19 pandemic, political decision makers are turning to mathematical models for advise [10]. A powerful quantitative concept to characterize the contagiousness and transmissibility of the new coronavirus is the basic reproduction number *R*_0_ [50]. This number explains–in simple terms–how many new infections are caused by a single one infectious individual in an otherwise completely susceptible population [13]. However, against many false claims, the *basic* reproduction number does not measure the effects of public health interventions [12]. Here, we quantify these effects, for every point in time, for every country, using the *effective* reproduction number *R*(*t*), a time-dependent metric that changes dynamically in response to community mitigation strategies and political actions. We learn the effective reproduction number from case data of the COVID-19 outbreak across Europe using Bayesian inference and systematically correlate it to political interventions.

### The classical SEIR model can predict a natural equilibrium and herd immunity

The SEIR model has advanced to the model of choice for the outbreak dynamics of COVID-19 [36]. It belongs to a class of infectious disease models that epidemiologists characterize as compartment models [14]. Compartment models represent the population via a sequence of compartments through which the population passes as the disease progresses. Out of the many different compartment models, the SEIR model seems best suited to mimic the epidemiology of COVID-19 via four compartments: the susceptible, exposed, infectious, and recovered populations. For more than three decades [6], epidemiologists have successfully applied the SEIR model to understand the outbreak dynamics of the measles, chickenpox, mumps, polio, rubella, pertussis, and smallpox [25]. For this class of diseases, the outbreak ends as the number of daily new cases, *β SI*, decreases. As such, the classical SEIR model is self-regulating: It naturally converges to an endemic equilibrium, at which either the susceptible group *S*, or the infectious group *I*, or both have become small enough to prevent new infections [32]. In epidemiology, this equilibrium is known as herd immunity [22]. In a homogeneous, well-mixed population, herd immunity occurs once a fraction of (1 −1/*R*_0_) of the population has become immune, either through the disease itself or through vaccination, see Appendix. For the basic reproduction number of *R*_0_ = 4.22±1.69 we found in this study, the herd immunity threshold would be 78%. This value is lower than 94% for the measles, 89% for chickenpox with, 86% for mumps and rubella, and 80% for polio [3], but significantly higher than the values of 16% to 27% for the seasonal flu [7]. The countries with the highest prevalence, Luxembourg with 0.72%, Sweden with 0.71%, and Spain with 0.64% [15], do currently not even come close to these values, not even when including asymptomatic cases that are believed to increase the prevalence by an order of magnitude [43], resulting in 7.2%, 7.1%, and 6.4%. Knowing the precise basic reproduction number of COVID-19 will be critical to estimate the conditions for herd immunity and predict the success of vaccination strategies.

### The dynamic SEIR model can predict the effects of public health interventions

The classical SEIR model is a valuable tool to understand the interplay of the susceptible, exposed, infectious, and recovered populations under unconstrained conditions. However, for the current COVID-19 pandemic, similar to SARS, MERS, or Ebola, the dynamics of these four populations are tightly regulated by public health interventions [10] including isolation, quarantine, physical distancing, and community containment [9, 53]. This implies that model parameters like the contact rate *β*, the rate at which an infectious individual comes into contact and infects others, are not constant, but modulated by social behavior and political action [5]. Here we explicitly account for a dynamic contact rate *β* (*t*) and express it as a function of the time-varying effective reproduction number *R*(*t*) [55]. This allows us to “bend the curve” and predict temporary equilibrium states, far away from the equilibrium state of herd immunity, but stable under current conditions [32]. Yet, these states can quickly become unstable again once the current regulations change [53]. Our dynamic SEIR model allows us to study precisely these scenarios.

### The time-varying effective reproduction number reflects the strength of public health interventions

To model temporal changes in the reproduction number, we propose a hyperbolic tangent type ansatz for the effective reproduction number *R*(*t*). This functional form can naturally capture the basic reproduction number *R*_0_, the converged reproduction number under the current constraints *R*_t_, the adaptation time *t*^∗^, and the transition time *T*, see Appendix. Figure 11 illustrates how our hyperbolic tangent type model compares against a constant and a random walk type reproduction number. The constant reproduction number in Figure 11, left, nicely captures the exponential increase during the early stages of the outbreak, but fails to “bend the curve” before herd immunity occurs. Nonetheless, several recent studies have successfully used an SEIR model with a constant reproduction number to model the outbreak dynamics of COVID-19 in China [42] and in Europe [34] by explicitly reducing the total population *N* to an affected population *N*^∗^ = *η N*. The scaling coefficient *η* = *N*^∗^/*N* is essentially a fitting parameter that indirectly quantifies the level of confinement [5]. For example, when averaged over 30 Chinese provinces, the mean affected population was *η* = 5.19 · 10^−5^±2.23±10^−4^, suggesting that the effect of COVID-19 was confined to only a very small fraction of the total population [42]. The Gaussian random walk in Figure 11, left, naturally captures the effects of public health interventions, however, in a daily varying, rather unpredictable way. It is a valuable method to analyze case data retrospectively, but since it does not allow for a closed functional form, it is not very useful to make informed predictions. We conclude that the hyperbolic tangent based ansatz in Figure 11, middle, with four physically meaningful parameters, is the most useful approach to represent the time-varying effective reproduction number *R*(*t*) for our current purposes.

**Fig 8.**
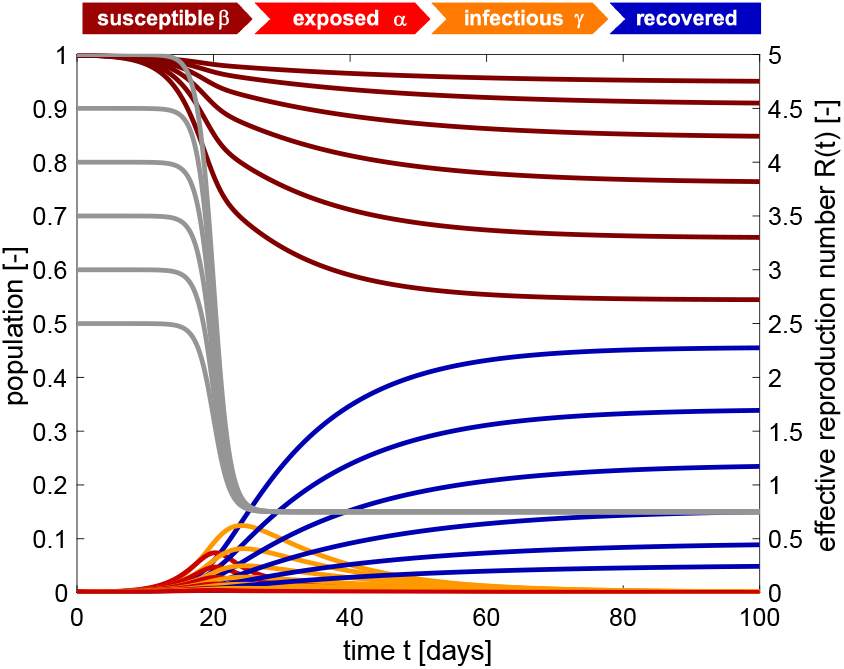
SEIR model with time-varying effective reproduction number. Increasing the basic reproduction number *R*_0_ increases the initial growth, and with it the number of cases. The temporary equilibrium for the smaller basic reproduction number of *R*_0_ = 2.5 is 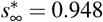 and 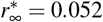 and for the larger basic reproduction number of *R*_0_ = 5.0 is 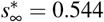 and 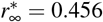. Latent period A = 2.5 days, infectious period *C* = 6.5 days, basic reproduction number *R*_0_ = [5.0.4.5.4.0,3.5,3.0.2.5], effective reproduction number *R*_*t*_ = 0.75, adaptation time *t*\* = 20 days, and transition time *T* = 15 days.

**Fig 9.**
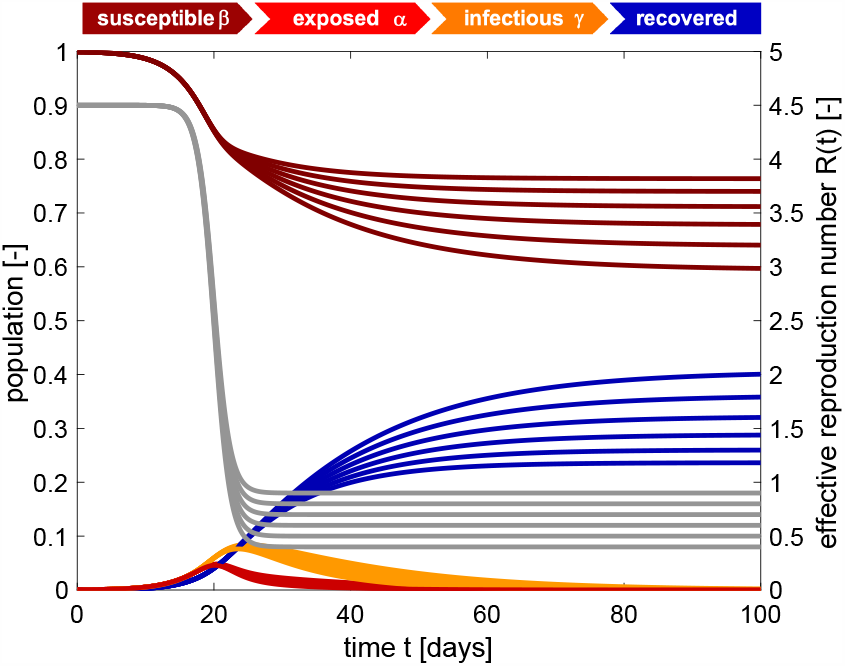
SEIR model with time-varying effective reproduction number. Increasing the reproduction number *R*_*t*_ decreases the effect of interventions and increases the number of cases. The temporary equilibrium for the smaller effective reproduction number of *R*_*t*_ = 0.4 is 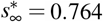 and 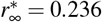 and for the larger effective reproduction number of *R*_0_ = 0.9 is 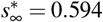 and 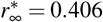. Latent period *A* = 2.5 days, infectious period *C* = 6.5 days, basic reproduction number *R*o = 4.5, effective reproduction number *R*_*t*_ = [0.4,0.5,0.6,0.7,0.8,0.9], adaptation time *t*\* = 20 days, and transition time *T* = 15 days.

**Fig 10.**
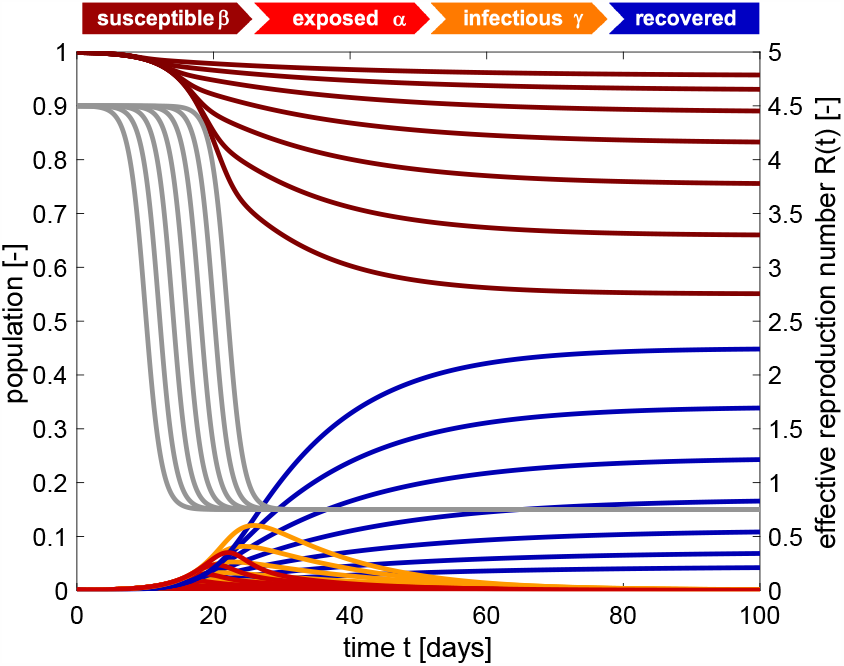
SEIR model with time-varying effective reproduction number. Increasing the adaptation time *t*\* to interventions increases the time spent at a high reproduction number, and with it the number of cases. The temporary equilibrium for the faster adaptation of *t*\* = 10 days is 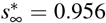 and 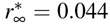 and for the slower adaptation of *t*\* = 22 days is 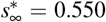 and 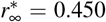. Latent period *A* = 2.5 days, infectious period *C* = 6.5 days, basic reproduction number *R*_0_ = 4.5, effective reproduction number *R*_*t*_ = *R*_0_/6 = 0.75, adaptation time *t*\* = [10,12,14,16,18,20,22] days, and transition time *T* = 15 days.

**Fig 11.**
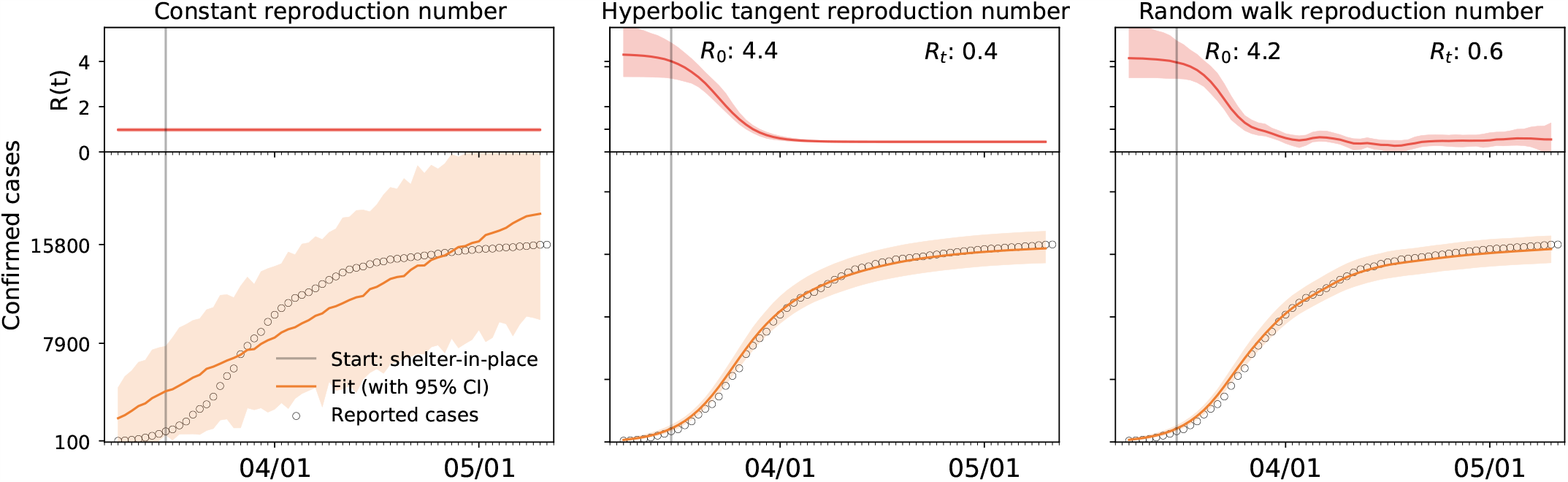
Time-varying effective reproduction number R(t). Comparison of constant, hyperbolic tangent, and random walk type ansatz. The constant effective reproduction number predicts an exponential increase in the number of cases that fits the initial but not for the later stages of the COVID-19 outbreak, left. The hyperbolic tangent type reproduction number predicts a smooth early increase and later saturation of the number of cases, middle. The random walk type reproduction number predicts a daily varying, non-smooth early increase and later saturation of the number of cases, right. Dots represent reported cases; orange curves illustrate fit with 95% confidence interval; red curves shows effective reproduction number with 95% confidence interval; here illustrated for the case of Austria.

### Bayesian inference identifies basic and effective reproduction numbers from reported cases

Unfortunately, we can neither measure the basic nor the effective reproduction number directly. However, throughout the past six months, the COVID-19 pandemic has probably generated more quantitative data than any infectious disease in history. Parametric Bayesian methods offers incredible opportunities to evaluate these data and learn correlations and trends [39]. Here we learn the effective reproduction number *R*(*t*) directly from the reported COVID-19 cases in all 27 countries of the European Union, starting from the day of the first reported case on January 24, until May 10, 2020. This not only allows us to identify the model parameters and confidence intervals, but also to quantify correlations between travel restrictions and reduced effective reproduction numbers. Table 2 and Figures 2 and 3 summarize our basic reproduction numbers *R*_0_ and effective reproduction numbers *R*_t_ for all 27 countries. Our mean basic reproduction number of *R*_0_ = 4.22±1.69 exceeds the first estimates of 1.4 to 2.5 from the World Health Organization based on a tracing study that reported a value of 2.2 during the early outbreak in Wuhan [33]. However, our results agree well with the more recent values of 5.7 for the Wuhan outbreak [47] and with a recent review that suggested values from 4.1 to 6.5 calculated with SEIR models [36]. Our basic reproduction number of 4.22 is lower than the numbers of 18 for measles, 9 for chickenpox, 7 for mumps, 7 for rubella, and 5 for poliomyelitis [3]. Compared to the SARS coronavirus with a range from 2 to 5 [36], our values of SARS-CoV-2 in Table 2 are rather on the high end, suggesting that the new coronavirus would spread more rapidly than SARS [54]. Knowing the precise basic reproduction number is critical to estimate the number of contacts to trace, if we want to successfully control the dynamics of COVID-19 through contact trancing [24].

### Political mitigation strategies reduce the effective reproduction number with a time delay of two weeks

Freedom of movement is the fundamental principle of the European Union. On March 13, 2020, the World Health Organization declared Europe the epicenter of the COVID-19 pandemic with more reported cases and deaths than the rest of the world combined [51]. To prevent a further spreading of the pandemic, four days later, for the first time in history, the European Union closed all its external borders [16]. In the following two weeks, the local governments augmented the European regulations with local lockdowns and national travel restrictions. Figure 4 shows that these measures had an enormous effect on the mobility within the European Union: By March 22, 2020, the average passenger air travel in Europe was cut in half, and as of May 10, it is reduced by 86% in Germany, 92% in France, 93% in Italy, and 95% in Spain [18]. These drastic actions have triggered an ongoing debate about the effectiveness of different outbreak strategies and the appropriate level of constraints [38]. Table 2 and Figures 4 to 7 summarize our time-varying effective reproduction number *R*(*t*) and highlight the time delay of its reduction with respect to the European travel restrictions. An important socio-economical metric is mean time delay of *Δt* = 17.24±2.00 days between the reduction of air traffic, driving, walking, and transit mobility and the inflection point of the reproduction number curve. Figures 5 and 7 show that this time delay varies hugely across Europe with the fastest response of 0.75 days in the Netherlands, followed by Germany with 3.25 days, Belgium with 4.00 days, and Italy with 5.00 days. These fast response times naturally also reflect decisions on the national level. France had the first reported COVID-19 case in Europe on January 24, 2020 and acted rigorously and promptly by introducing the first national measures on March 16 [52]. Similarly, Italy, Spain, and Germany had introduced their national measures on March 9, March 9, and March 13, 2020 [48]. Figures 5 and 7 clearly highlight the special role of Sweden, where the government focusses efforts on encouraging the right behavior and creating social norms rather than mandatory restrictions: The time delay of 23.75 days is above the European Union average of 17.24 days, and Sweden is one of the few countries where the effective reproduction number has not yet decreased below one. Taken together, these results confirm that, especially during the early stages of an outbreak, controlling mobility can play a critical role in spreading a disease [8]. However, these drastic political measures have stimulated an active ongoing debate when and how it would be safe to lift these restrictions.

### Exit strategies will have different effects in individual countries

Political decision makers around the globe are currently trying to identify safe exit strategies from global travel restrictions and local lockdown. Mathematical models can provide guidelines and answer what-if scenarios. Our predictions in Figure 1 show projections of the number of total cases, for three possible exit strategies from lockdown: a continuation at a constant effective reproduction number *R*_t_, a gradual return to the basic reproduction number *R*_0_ within three months, and a rapid to *R*_0_ within one months. Naturally, the case numbers increase in all three cases, with the steepest increase for the most rapid return. Interestingly, our method provides significantly different confidence intervals for different countries suggesting that a controlled return will be more predictable in some countries like Austria and less in others. Our projections suggest that in Sweden, were policy makers had encouraged each individual to take responsibility for their own health rather than enforcing political constraints, the projected case numbers will follow the current curve, without major deviations. Strikingly, in most countries, the newly reported case numbers upon gradual reopening, from May 10 to June 20, 2020, follow the dashed brown curves of the prediction with a constant effective reproduction number. This suggests that most countries have learnt how to successfully control the pandemic and manage new outbreaks.

### Limitations

Just like any infectious disease model, our model inherently faces limitations associated with data uncertainties from differences in testing, inconsistent diagnostics, incomplete counting, and delayed reporting. For our specific study of COVID-19, we encounter a few additional limitations: First, although a massive amount of data are freely available through numerous well-documented public databases, the selection of the model naturally limits what we can predict and it remains challenging to map the available information into the format of the SEIR model. Second, the initial conditions for our exposed and infectious populations will always remain unknown and many new first cases have been reported throughout the past couple of weeks. To reduce the influence of unknown initial conditions, our parametric Bayesian inference algorithm learns these populations alongside the effective reproduction number. Third, in its current state, our model does not distinguish between community mitigation strategies, local public health recommendations, and global political actions [9]. We are currently integrating the current approach into a global network model that will provide more granularity to include other community mitigation strategies in addition to mobility. Fourth, our current model is not directly informed by mobility data. We have recently proposed a new method that uses a stochastic process to directly incorporate mobility as a latent variable into the present SEIR model framework [35]. Fifth, and probably most importantly, our current knowledge limits our ability to make firm predictions about the recovered group, which will be critical to estimate the return to normal. Recent studies have shown that the unreported asymptomatic population is huge, up to an order of magnitude larger than the reported symptomatic population traced in our study [43]. A related challenge is that the number of reported cases strongly depends on the testing strategy of each country. A possibility to eliminate testing bias could be to use death counts rather than case counts [23]; however, this would also require a consistent Europe-wide definition of death with versus death caused by COVID-19. In general, more targeted tests will be needed to identify the size of the asymptomatic population and explore whether it behaves differently in terms of contact rate and infectious period, which would both radically change the overall reproduction number. As more data become available, we are confident that we will learn from uncertainty quantification, become more confident in our model predictions, and learn how to quickly extract important trends.

## 5 Conclusion

We quantified the effectiveness of public health interventions using the effective reproduction number *R*, the time-varying reproduction number of the COVID-19 pandemic, across all 27 countries of the European Union. We adopted an SEIR epidemiology model with a dynamic effective reproduction number, which we learned for each country from its individual reported cases using Bayesian inference. We found that, during the early stages of the COVID-19 outbreak, the basic reproduction number across Europe was *R*_0_ = 4.22±1.69. Massive public health interventions as well as social learning have successfully reduced the effective reproduction number to *R*_t_ = 0.67±0.18 by May 10, 2020. Strikingly, this reduction displays a strong correlation with mobility in the form of air travel, driving, walking and transit mobility with a mean time delay of 17.24±2.00 days. This time delay is an important metric as we seek to identify safe exit strategies from current lockdown and travel restrictions. To highlight the predictive potential of our model, we simulated different exit strategies from lockdown that either maintain the current status quo, gradually return to normal, or rapidly return to the early exponential growth. Upon gradual reopening, from May 10 to June 20, 2020, the newly reported case numbers in most countries followed the prediction that maintained the current effective reproduction number suggesting that most countries were able to successfully manage the pandemic and control new outbreaks. Our dynamic epidemiology model provides the flexibility to simulate the effects and timelines of various outbreak control and exit strategies to inform political decision making and identify solutions that minimize the impact of COVID-19 on global health.

## Appendix

### The SEIR model

The SEIR model is a popular model in the epidemiology of infectious diseases [25]. It represents the timeline of a disease through four compartments that characterize the dynamics of the susceptible, exposed, infectious, and recovered populations [6]. The transition between these populations is governed by a set of ordinary differential equations [29],

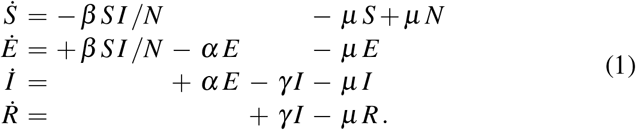

The transition rates between the four populations, the contact rate *β*, the latency rate *α*, and the infectious rate *γ*, are inverses of the contact period *B* = 1/*β*, the latent period *A* = 1/*α*, and the infectious period *C* = 1/*γ*. The set of equations (1) includes vital dynamics at an equivalent birth and death rate *µ*, such that the sum of all four equations, (1.1) to (1.4), is equal to zero,

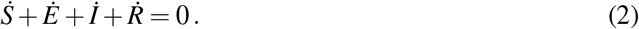

This implies that the sum of the four populations is constant and equal to the total population *N*,

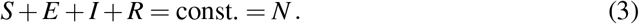

For the SEIR model with vital dynamics (1), the basic reproduction number *R*_0_, the number of new infections caused by one infectious individual in a completely susceptible population [13], is

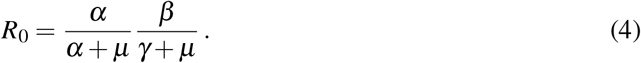

The magnitude of *R*_0_ plays a critical role in the outbreak dynamics of an infectious disease [36]. Here we are interested in studying the outbreak dynamics of COVID-19, for which the time period is short, and we can neglect the effects of vital dynamics. This implies that the set of equations (1) reduces to the following system,

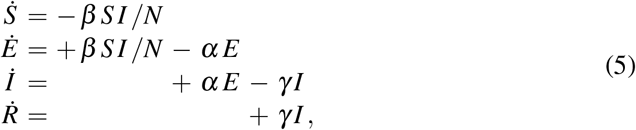

and the basic reproduction number (4) simplifies to the following expression,

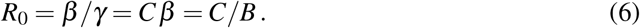

Many infections diseases, including COVID-19, display a significant latent period during which individuals have been infected but are not yet infectious themselves. These individuals are represented through the exposed population *E*. A special case of the SEIR model is the SIR model, which follows from the set of equations (5) with *α →* ∞ as

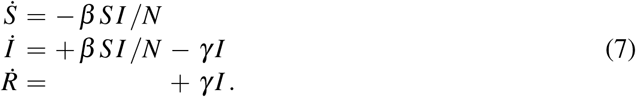

While the SIR model is conceptually simpler and lends itself to closed form solutions, for the outbreak dynamics of the COVID-19 pandemic, the invisible exposed, but not yet infectious population plays a critical role. Throughout this study, we therefore focus on the SEIR model. We reparameterize the absolute SEIR model (5) and scale it by the total population *N*, to obtain the fractions of the susceptible, exposed, infectious, and recovered populations,

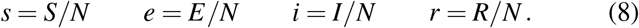

This introduces the relative SEIR model,

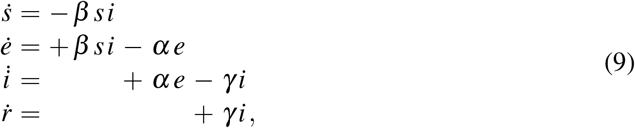

parameterized in the fractional populations, *s, e, i*, and *r*, which sum up to one,

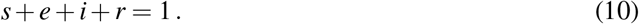

### Endemic equilibrium

The hallmark of a typical epidemic outbreak is that it begins with a small infectious population *I*_0_. The infectious population *I*(*t*) increases, reaches a peak, and then decays to zero [25]. Throughout the outbreak, the susceptible population *S*(*t*) decreases, but the final susceptible population *S*_∞_ always remains larger than zero. This final state is called the endemic equilibrium. To estimate the endemic equilibrium of the COVID-19 pandemic, we divide equation (5.1) by equation (5.4),

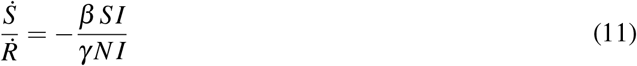

Separation of the variables and using the definition of the basic reproduction number *R*_0_ = *β*/*γ* yields the following equation,

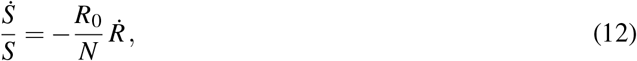

which we integrate in time,

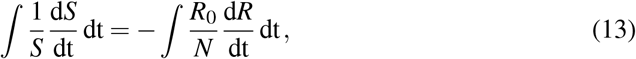

to obtain the following expression,

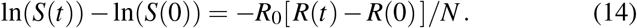

Here *S*(0) and *R*(0) are the initial susceptible and recovered populations and *S*(*t*) and *R*(*t*) are these populations at time *t*. Using ln(*S*(*t*)) −ln(*S*(0)) = ln(*S*(*t*)/*S*(0)) and applying the exponential function on both sides of the equation introduces the following explicit representation for the susceptible population at time *t*,

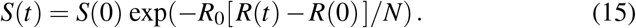

According to equation (8), we scale the populations with the total population *N* as *s*_0_ = *S*(0)/*N* and *r*_0_ = *R*(0)/*N*, and evaluate equation (15) at the limit *t→* ∞ with *s*_∞_ = *S*(∞)/*N, e*_∞_ = 0, *i*_∞_ = 0, and *r*_∞_ = *R*(∞)/*N* = 1− *s*_∞_, to obtain the following expression for the susceptible population at endemic equilibrium,

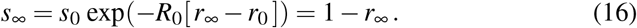

This transcendental equation has an explicit solution in terms of the Lambert function *W*,

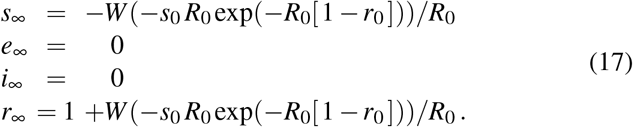

The endemic equilibrium condition (17) confirms that, unless *S*(0) = 0, the final susceptible population will always be larger than zero, *S*_∞_ > 0 [32].

### Public health interventions

The classical SEIR model (1) assumes that the disease develops freely and that the contact rate *β*, latency rate *α*, and infectious rate *γ* are constant throughout the course of the outbreak. It is obvious that the contact rate *β* will change in response to community mitigation strategies and political actions, e.g., local lockdown and global travel restrictions [20]. Here, to account for the effects of public health interventions, we introduce a time-varying contact rate *β* (*t*) and rewrite the system of equations (5),

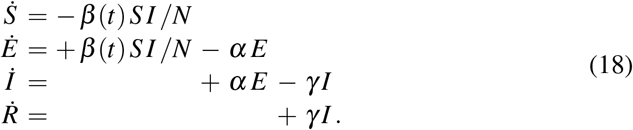

We make a hyperbolic tangent type ansatz for the contact rate *β* (*t*),

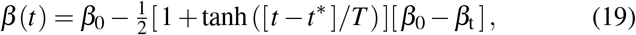

where *β*_0_ is the initial contact rate at the onset of the pandemic, *β*_*t*_ is the contact rate in response to public health interventions, *t*^∗^ is the adaptation time, and *T* is the transition time. For easier interpretation, we reparameterize the system (18) in term of the time-dependent effective reproduction number *R*(*t*) = *β* (*t*)/*γ*,

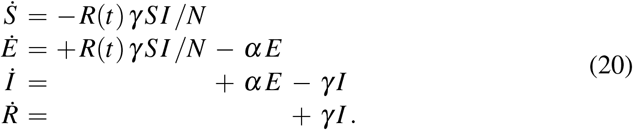

With equation (19), the effective reproduction number takes the following hyperbolic tangent type form,

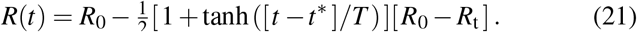

This ansatz ensures a smooth transition from the initial basic reproduction number *R*_0_ = *β*_0_/*γ* at the beginning of the outbreak to the effective reproduction number *R*_t_ = *β*_t_/*γ* in response to public health interventions, where *t*^∗^ and *T* are the adaptation and transition times. From equation (16), we can estimate the constrained equilibrium in response to public health interventions,

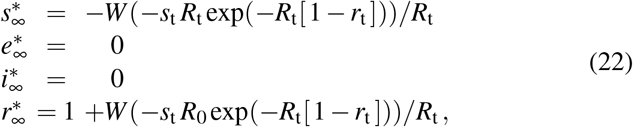

where *s*_t_ = *s*_*t*_∗+*T*/2 and *r*_t_ = *r*_*t*_∗+*T*/2 are the fractions of the susceptible and recovered populations at time *t* = *t*^∗^ + *T*/2, the time at which the effective reproduction number has fully adopted the new value *R*(*t*) = *R*_t_. Importantly, this constrained equilibrium is not equivalent to the natural endemic equilibrium, 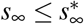 and 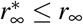, since *R*_t_ ≤ *R*_0_.

### Time-varying effective reproduction number

Figures 8 to 10 illustrate the outbreak dynamics of our SEIR model with a time-varying effective reproduction number. The gray curves highlight the hyperbolic tangent type nature of the effective reproduction number *R*(*t*), the dark red, red, orange, and blue curves illustrate the dynamics of the susceptible *S*, exposed *E*, infectious *I*, and recovered *R* populations. Unless stated otherwise, we use a latent period of *A* = 2.5 days, an infectious period of *C* = 6.5 days, a basic reproduction number of *R*_0_ = 4.5, a reproduction number under public health interventions of *R*_t_ = 0.75, and adaptation and transition times of *t*^∗^ = 20 days and *T* = 15 days. In all simulations, the effective reproduction number *R*(*t*) transitions gradually from the initial basic reproduction number *R*_0_ at the beginning of the outbreak to the effective reproduction number *R*_t_ associated with the public health interventions. The adaptation time *t*^∗^ marks the midpoint of the transition and the transition time *T* is its duration. The outbreak is more pronounced for larger basic reproduction numbers *R*_o_ as we see in Figure 8, for larger intervention related reproduction numbers *R*_*t*_ as we see in Figure 9, and for larger adaptation times *t*^∗^ as we see in Figure 10.

### Constant, hyperbolic tangent, and random walk type effective reproduction numbers

To illustrate the effect of different time-varying effective reproduction numbers, we compare three different methods: a constant effective reproduction number, a smoothly decaying effective reproduction number of hyperbolic tangent type, and a daily varying effective reproduction number that follows a Gaussian random walk. The constant reproduction number has one parameter *R*_*t*_ = *R*_0_. The hyperbolic tangent type reproduction number. 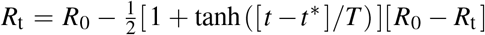, has four parameters, the basic and effective reproduction numbers *R*_0_ and *R*_*t*_, the adaptation time *t*\*, and the time delay *Δt*. The Gaussian random walk has three parameters, the drift *μ*, the daily stepwidth τ = τ_1_ /[1.0 — *s*], and the smoothing parameter s. Table 3 summarizes the prior distributions for all three methods. Figure 11 compares the constant, hyperbolic tangent, and random walk type effective reproduction numbers for the example of Austria. The three graphs illustrate the number of reported cases as dots, the model fit as orange curves with 95% confidence interval, and the effective reproduction numbers as red curves with 95% confidence interval. Of all three methods, the constant ansatz can fit the early exponential increase of the COVID-19 outbreak, but not the later saturation. The random walk type ansatz can fit both the early exponential increase and the later saturation, but not with a closed form expression. Only the hyperbolic tangent type ansatz provides both a good fit and a closed functional form to compare the time lines of the outbreak in different countries and make informed predictions.

**Table 3.**
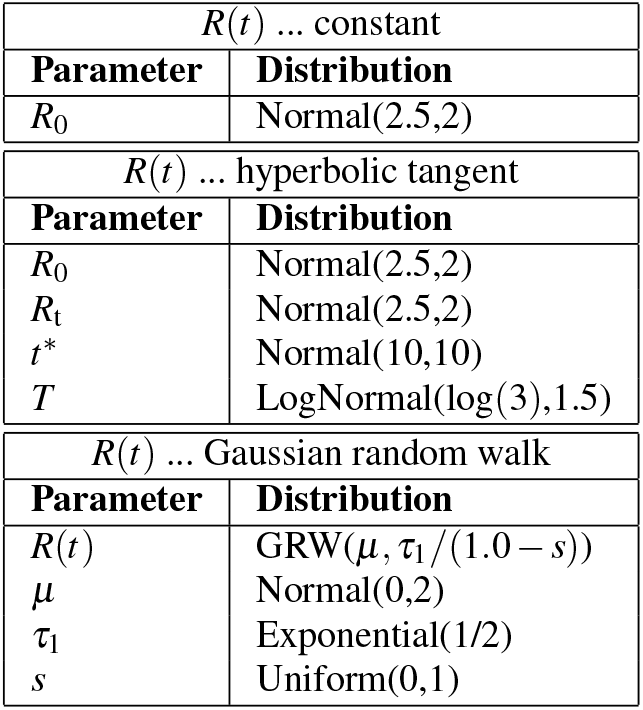
Prior distributions for time-varying effective reproduction number R(t) of constant, hyperbolic tangent, and Gaussian random walk type.

| $R(t)$ ... constant | |
| --- | --- |
| Parameter | Distribution |
| $R_0$ | Normal(2.5,2) |
| $R(t)$ ... hyperbolic tangent | |
| Parameter | Distribution |
| $R_0$ | Normal(2.5,2) |
| $R_t$ | Normal(2.5,2) |
| $t^*$ | Normal(10,10) |
| $T$ | LogNormal(log(3),1.5) |
| $R(t)$ ... Gaussian random walk | |
| Parameter | Distribution |
| $R(t)$ | GRW($\mu, \tau_1 / (1.0 - s)$) |
| $\mu$ | Normal(0,2) |
| $\tau_1$ | Exponential(1/2) |
| $s$ | Uniform(0,1) |

### Herd immunity

An important consequence of the basic reproduction number *R*_0_ is the condition for herd immunity [12]. Herd immunity occurs once the immune population, in our case the recovered population *R*, is large enough to protect susceptible individuals from infection [22]. We can express herd immunity in terms of the recovered fraction *r*, or in terms of the absolute recovered population *R*,

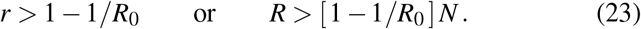

Importantly, upon relaxing public health interventions, the condition for herd immunity is not *R* > 1 −1/*R*_t_. Herd immunity is not a function of the reproduction number under public health interventions *R*_t_–which is usually much smaller than the basic reproduction number *R*_0_–but will depend on the natural basic reproduction number *R*_0_ under unconstrained conditions.

## Data Availability

All data are publicly available in public databases with references listed in this article.

## Acknowledgments

We acknowledge stimulating discussions with Dr. Francisco Sahli Costabal. This work was supported by a DAAD Fellowship to Kevin Linka and a Stanford Bio-X IIP seed grant to Mathias Peirlinck and Ellen Kuhl and by the National Institutes of Health Grant U01 HL119578.

## Notes

### Competing Interest Statement

The authors have declared no competing interest.

